# Does weather affect the growth rate of COVID-19, a study to comprehend transmission dynamics on human health

**DOI:** 10.1101/2020.04.29.20085795

**Authors:** Arjun Sil, Vanapalli Naveen Kumar

## Abstract

The undefendable outbreak of novel coronavirus (SARS-COV-2) lead to a global health emergency due to its higher transmission rate and longer symptomatic duration, created a health surge in a short time. Since Nov 2019 the outbreak in China, the virus is spreading exponentially everywhere. The current study focuses on the relationship between environmental parameters and the growth rate of COVID-19. The statistical analysis suggests that the temperature changes retarded the growth rate and found that −6.28°C and +14.51°C temperature is the favorable range for COVID-19 growth. Gutenberg-Richter’s relationship is used to estimate the mean daily rate of exceedance of confirmed cases concerning the change in temperature. Temperature is the most influential parameter that reduces the growth at the rate of 13–16 cases/day with a 1°C rise in temperature.

## Introduction

Several virus-infected diseases occur seasonally on a regular basis affecting human health every year. A virus invades the human body by hijacking the internal cell machinery to introduce its own genetic material in order to reproduce more virus particles in the host. Since ancient times, numerous deaths including both humans and animals occurred due to several seasonal virus/bacterial infections to which people used to give their effort for survival by minimizing this infection through inventing various vaccines or medicines. However, so far already the deadly Covid-19 virus originated diseases created pandemic that has affected a more significant part of the world population.

Currently, the whole world is struggling to sustain the menacing health emergency due to the outbreak of novel coronavirus Sars-Cov-2, causing COVID-19. Sars-Cov-2 is the 7th human pathogen recognized in the coronavirus family (M. Ceccarelli and M. Berretta, 2020) and is also similar to the Sars-Cov in 2002, Sars-Cov affected people globally with around 10% mortality, whereas MERS in 2012 caused 34% death among the infected people. It is reported (Li et al. 2020) that SARS-Cov-2 has a reproductive number (R_0_) of 2.2 indicating that a patient on average spreads the virus to 2.2 people around him. The study also suggests that the average incubation period is around 5.2 days, with influenza doubled in size for every 7.4 days as the host transmits to close contacts before exhiniting symptoms (within 14 days), leading to an exponential chain transmission. The Covid-19 mortality rate as on 31^st^ March 2020 is found to be about 4.97%. However, the mortality rate of Covid-19 is less compared to the other two infectious diseases, nevertheless, the crucial fact is that it is infecting or transmitting exponentially from human to human creating a health surge in a short lapse of time. However, the challenge is to maintain R_0_<1 so that the spreading capability of Covid-19 is reduced and it could be attained through mitigation strategies such as managing social distance, using a mask and maintaining personal hygiene.

The recent study observes that the mortality rate of Covid-19 is higher for males than females i.e., 2.8% and 1.7% respectively. Further, On the other hand, considering the global scenario, the fatality rate of old people (>60 years of age) become profoundly affected with a mortality rate of 26.4%, whereas the people with age ranging between 10–59 years is 2.3 % and no death below 10 years is reported so far. However, around 56% of the global population (2019 census) live in urban areas and are highly vulnerable to the virus as WHO declared it, as a droplet oriented transmission, and it is difficult to contain the spread as maintaining social distancing is difficult in urban areas. The global location-wise data show exponential growth in infections as in figure 1. Indeed, the severely infected top 20 countries are presented in table 1.

**Table 1:** Top 20 countries severely affected are presented according to the order of infection cases

| Country | Confirmed Cases | Deaths | Cases as % of Global | Mortality Rate |
| --- | --- | --- | --- | --- |
| USA | 104837 | 1704 | 17.64 | 1.63 |
| Italy | 86498 | 9134 | 14.55 | 10.56 |
| Mainland China | 81394 | 3295 | 13.69 | 4.05 |
| Spain | 65719 | 5138 | 11.06 | 7.82 |
| Germany | 53340 | 395 | 8.97 | 0.74 |
| Iran | 32332 | 2378 | 5.44 | 7.35 |
| France | 29566 | 1696 | 4.97 | 5.74 |
| UK | 14543 | 759 | 2.45 | 5.22 |
| Switzerland | 12928 | 231 | 2.17 | 1.79 |
| South Korea | 9478 | 144 | 1.59 | 1.52 |
| Netherlands | 8603 | 546 | 1.45 | 6.35 |
| Austria | 7712 | 58 | 1.3 | 0.75 |
| Belgium | 7284 | 289 | 1.23 | 3.97 |
| Turkey | 5698 | 92 | 0.96 | 1.61 |
| Canada | 4757 | 55 | 0.8 | 1.16 |
| Portugal | 4268 | 76 | 0.72 | 1.78 |
| Norway | 3773 | 19 | 0.63 | 0.5 |
| Australia | 3635 | 14 | 0.61 | 0.39 |
| Brazil | 3477 | 93 | 0.58 | 2.67 |
| Israel | 3460 | 12 | 0.58 | 0.35 |
| Sweden | 3046 | 92 | 0.51 | 3.02 |
| Malaysia | 2161 | 26 | 0.36 | 1.2 |
| Denmark | 2154 | 41 | 0.36 | 1.9 |
| Czech Republic | 2062 | 9 | 0.35 | 0.44 |
| Ireland | 1819 | 19 | 0.31 | 1.04 |

**Figure 1:**
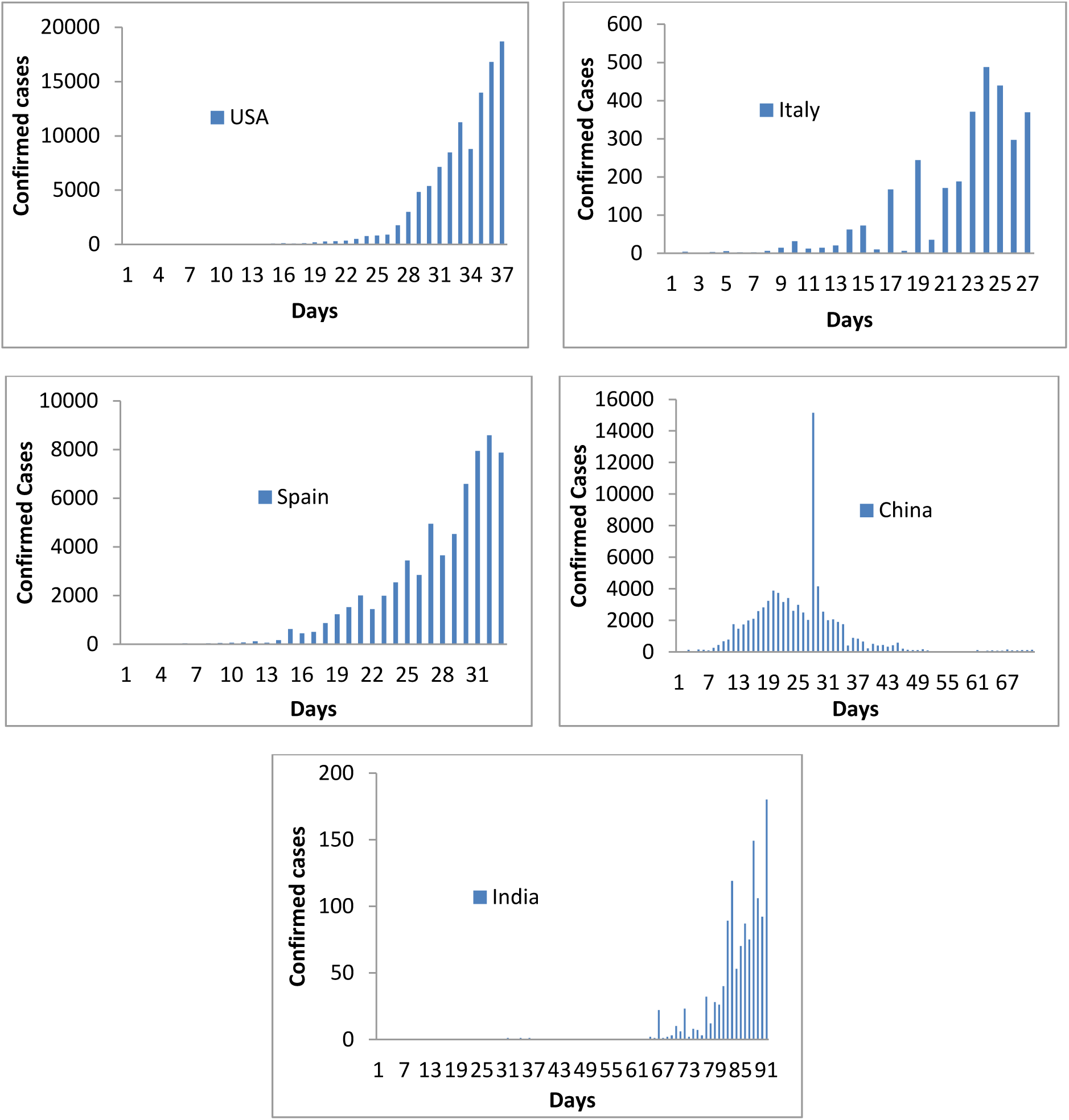
Frequency diagram of daily infected confirmed cases reported for top 4 countries including India a) USA, b) Itlay, c) Spain, d) China & e) India

The Covid-19 outbreak came into the spotlight at the end of December 2019 in the Wuhan city of China and within no time turned into pandemic through spreading over to most of the countries. However, globally till 31 March 2020, the WHO reported a total of 903540 confirmed infected cases and 47241 deaths (mortality rate 5.2%). In reality, the actual infected numbers may go much higher due to non reporting facts during the initial cases, or it is being detected slightly in later days. Further, these cases might go much higher within a short span. However, an appealing fact is that the number of reported cases are varying region to region or country to country with respective to their population. India is the 2^nd^ largest population in the world with 1370 million as per 2019 census data, however, the total reported cases and deaths till March 2020 are 2113 and 60 respectively, whereas in USA, Itlay, China and Spain, the total infected cases are 104837 (17.64%), 86498 (14.55%), 81394(13.69%), and 65719 (11.06%) respectively (Table 1). Further, the death rate (10.56%) in Itlay becomes higher compared to the USA (1.53%), China (4.05%) and India (2.8%).

## Environmental conditions

A study (Lin et al. 2006) states that Sars-Cov loses its stability and couldn’t facilitate higher transmission in tropical areas at a higher temperature and relative humidity. This evidentially emblazes an indication that like its predecessor, Sars-Cov 2 incidence may also be less at a higher temperature and humidity. However, a recent study (Araújo and Naimi, n.d.) that climatic conditions constrain the spread of the Sars-Cov-2. It is reported that the cold climates are more favorable for the spread of the virus rather than arid and tropical climates. Another study (Sajadi et al., n.d.) supports this discussion stating that the virus spread is more and have smiliar cluster patterns in the colder regions within 30–50°N having average temperatures between 5–11°C associated with low absolute humidity (4–7 g/m^3^). Further, another study (Wang et al. 2020) reports that a rise in 1°C temperatures leads to a 0.86 decrease in cumulative cases. However, another study (Gupta, n.d.) observed that a 1°C increase in temperature above 5°C reduces the transmission by 10%. These reported studies provide substantial evidence that the climatic conditions can regulate the transmission of the rapidly spreading novel coronavirus Sars-Cov-2.

Indeed, out of various environmental factors such as humidity, cloud cover, wind speed and precipitation, temperature contributes significantly to the decrease in the growth rate of infection. However, to study the effect of temperature, initially, a set of global temperature data has been collected for the past four months since November 2019 (source: University of Mine) as shown in Figure 2. It has been observed that all the highly affected countries USA, Itlay, Spain, and China are lying in between latitude 30–50°N from the equator and also the temperature gradient is below 15°C for the considered period. However, the temperature is high (> 15°C) in the regions lying between latitude +40°N to −40°S. The countries lying within this range are India, Africa, and Australia exhibiting a comparatively lower infection rate as well as the death toll among the top 20 list of countries. Some few cases are found in different scenarios with slightly higher infection rates such as Malaysia, Indonesia, and Australia. This may be due to higher topographical elevation or RL of the respective locations and also could be due to seasonal change in the location. This gives an indication that temperature plays a vital role in the outbreak or spreading of this virus.

**Figure 2:**
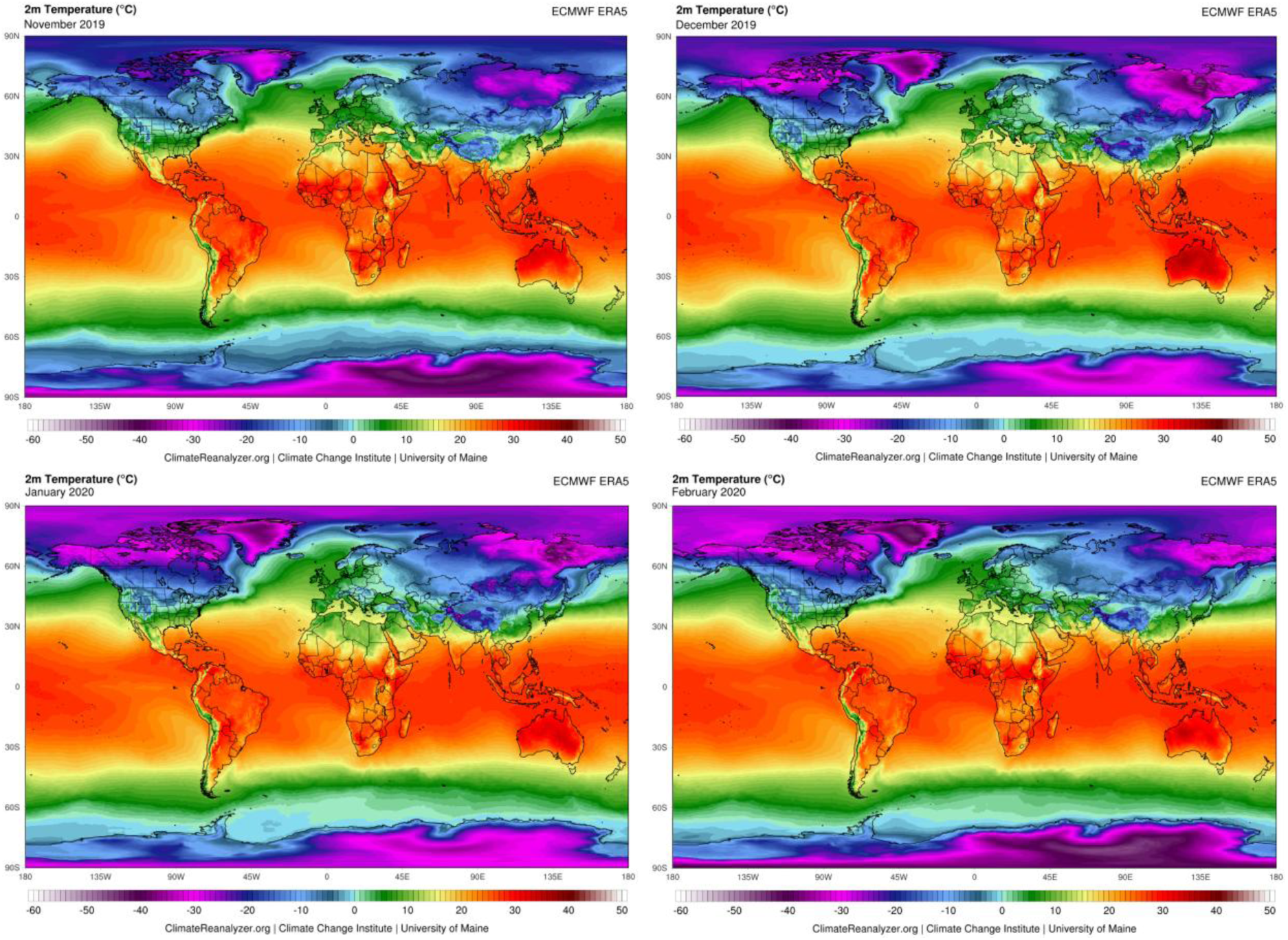
Global Temperatures variations for the last four months a) Nov 2019, b) Dec 2019, c) Jan 2020, d) Feb 2020. (source: University of Mine)

The next factor that contribute could be precipitation, however, in this regards the past five months data (Nov 2019 – March 2020) is carefully analyzed and presented in Figure 3. A higher precipitation is observed close to the equator. On the other hand, it has been observed that those highly affected regions such as USA, Europe, South China and Japan have no such significant precipitation on their land but their neighboring areas encountered moderately high rainfall (Figure 3). Therefore, it could be inferred that the neighboring regions having rainfall < 400 mm have significantly contributed to the favorable growth of Covid-19.

**Figure 3:**
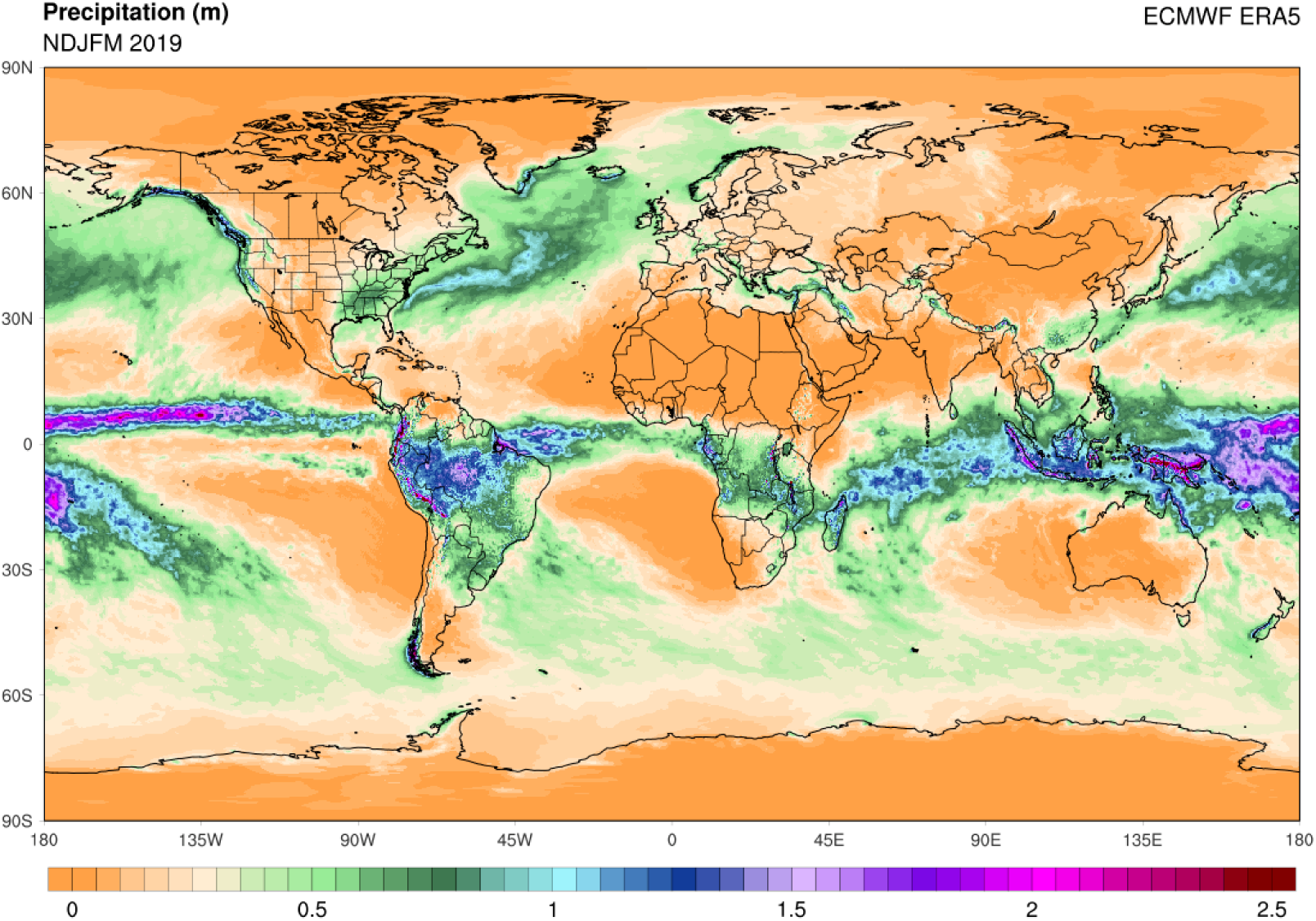
Variations of precipitation (m) worldwide during the last five months (Nov 2019-March 2020)

The third factor that could contribute is wind speed and the past five months wind data is shown in Figure 4. It has been observed from figure 4 that the speed of wind for the highly affected regions such as USA, Europe, and China experienced a velocity of about 10–12 m/s during the considered period. However, India, South America and central South Africa experienced a wind speed <2 m/s. This observation suggests that the wind speed may also contribute to favorable growth of Covid-19.

**Figure 4:**
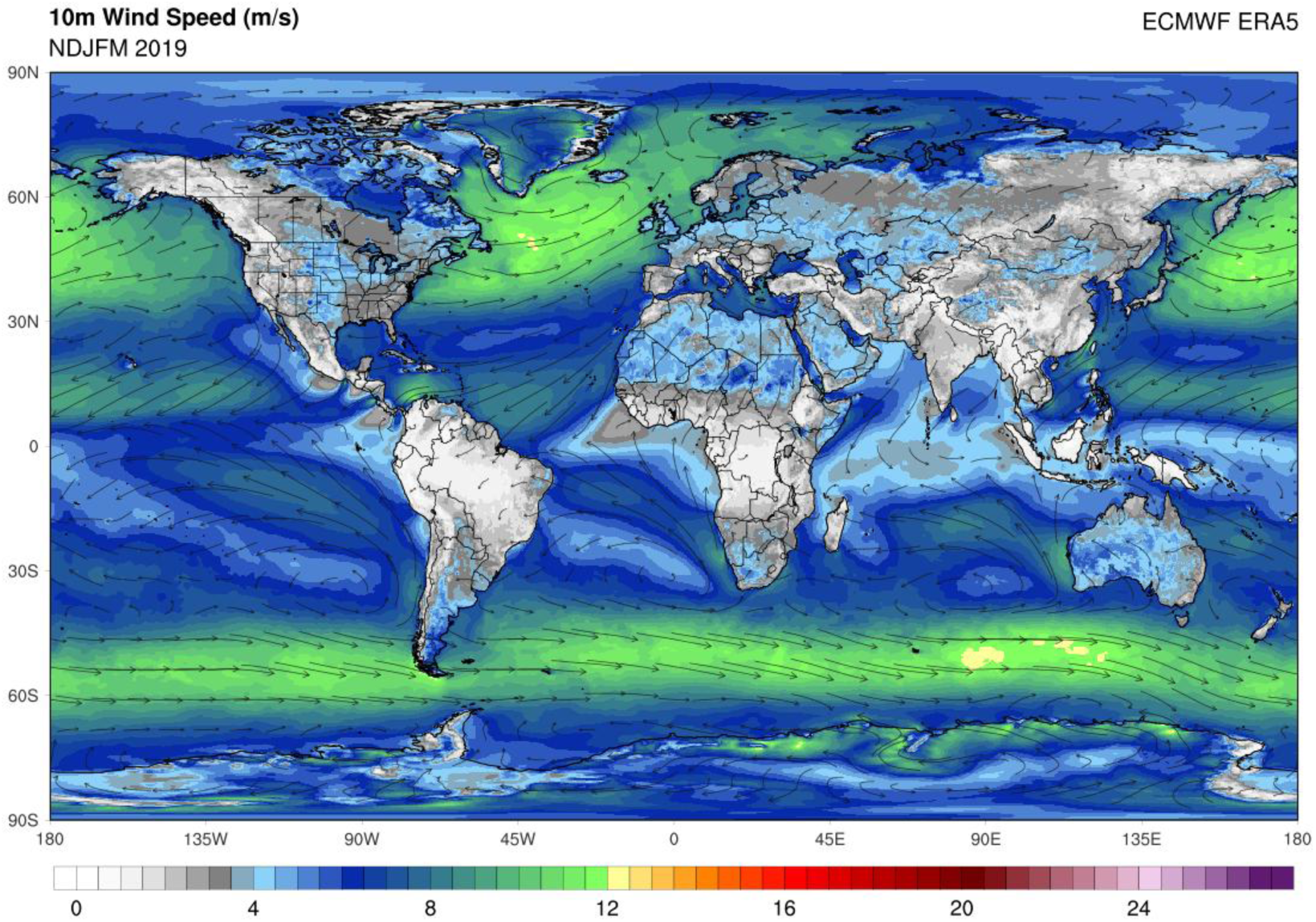
Variations of wind speed (m/s) worldwide during the last five months (Nov 2019-March 2020)

The Fourth factor that could contribute is a cloud cover. The higher cloud coverage is observed (Figure 5) slightly away from the equator. However, those highly affected countries have approximate cloud coverage ranging between 70–80% in these past four months.

**Figure 5:**
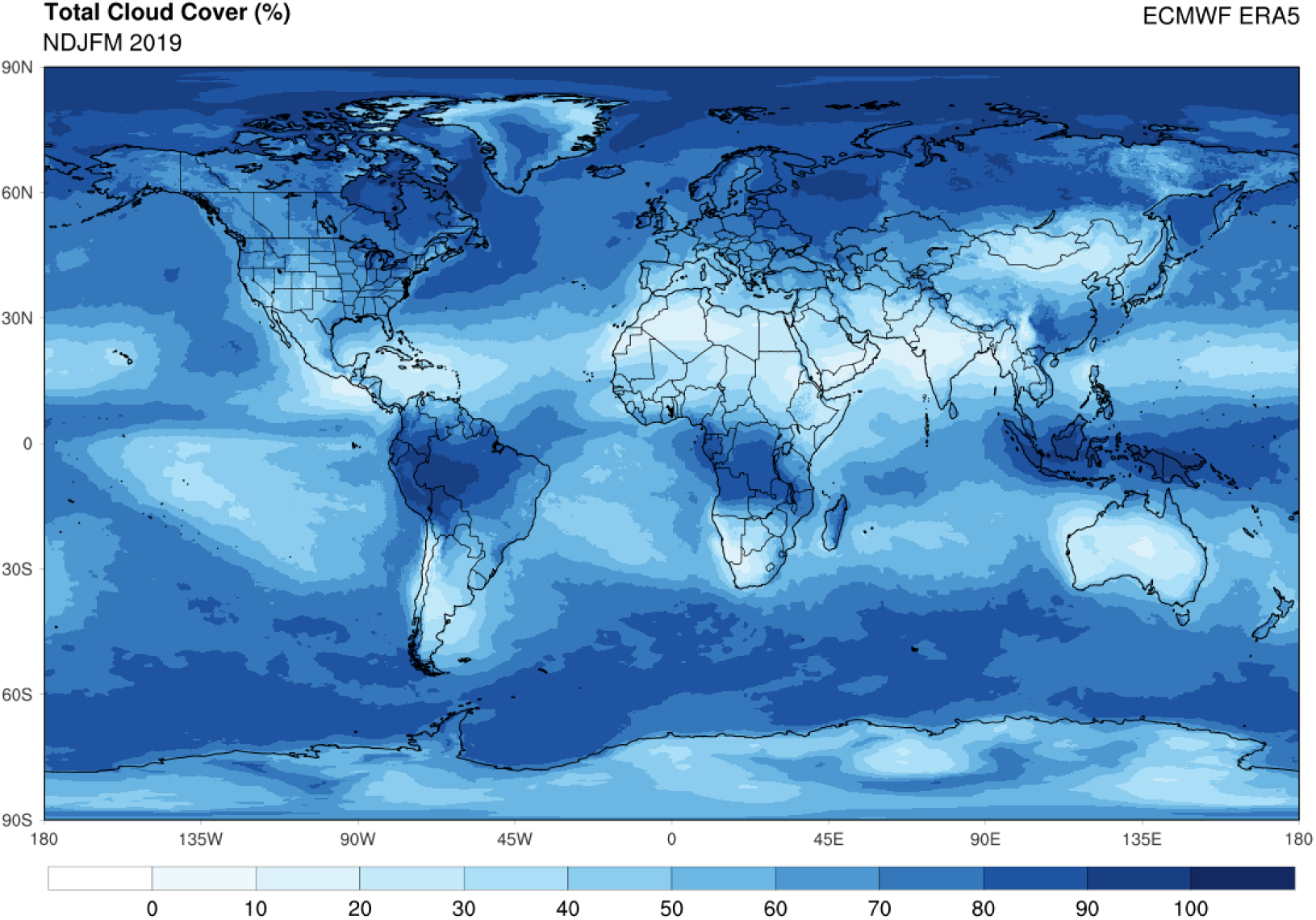
Variations of cloud coverage (%) worldwide during the last five months (Nov 2019-March 2020)

## Data Collection and processing

To study the effects of temperature on the growth of Covid-19, the necessary data is extracted from various reliable sources such as Wang et al (2020) and the University of Mine. However, the Wang et al (2020) presented the data from Jan 2020 to Feb 4, 2020 considering the confirmed cases observed for all the cities or regions worldwide and reported the daily avg, min and max temperatures for the respective regions. This extracted data is considered for the present study combining with other data statistically to examine the temperature dependency on the growth of Covid-19. Further, considering the spatial variations of temperature the regions under study USA, Europe, China and Canada, Siberia, Africa, India are classified as high and least affected areas respectively. The collected regional mean monthly temperature data for the last 41 years is presented in Figure 6.

**Figure 6:**
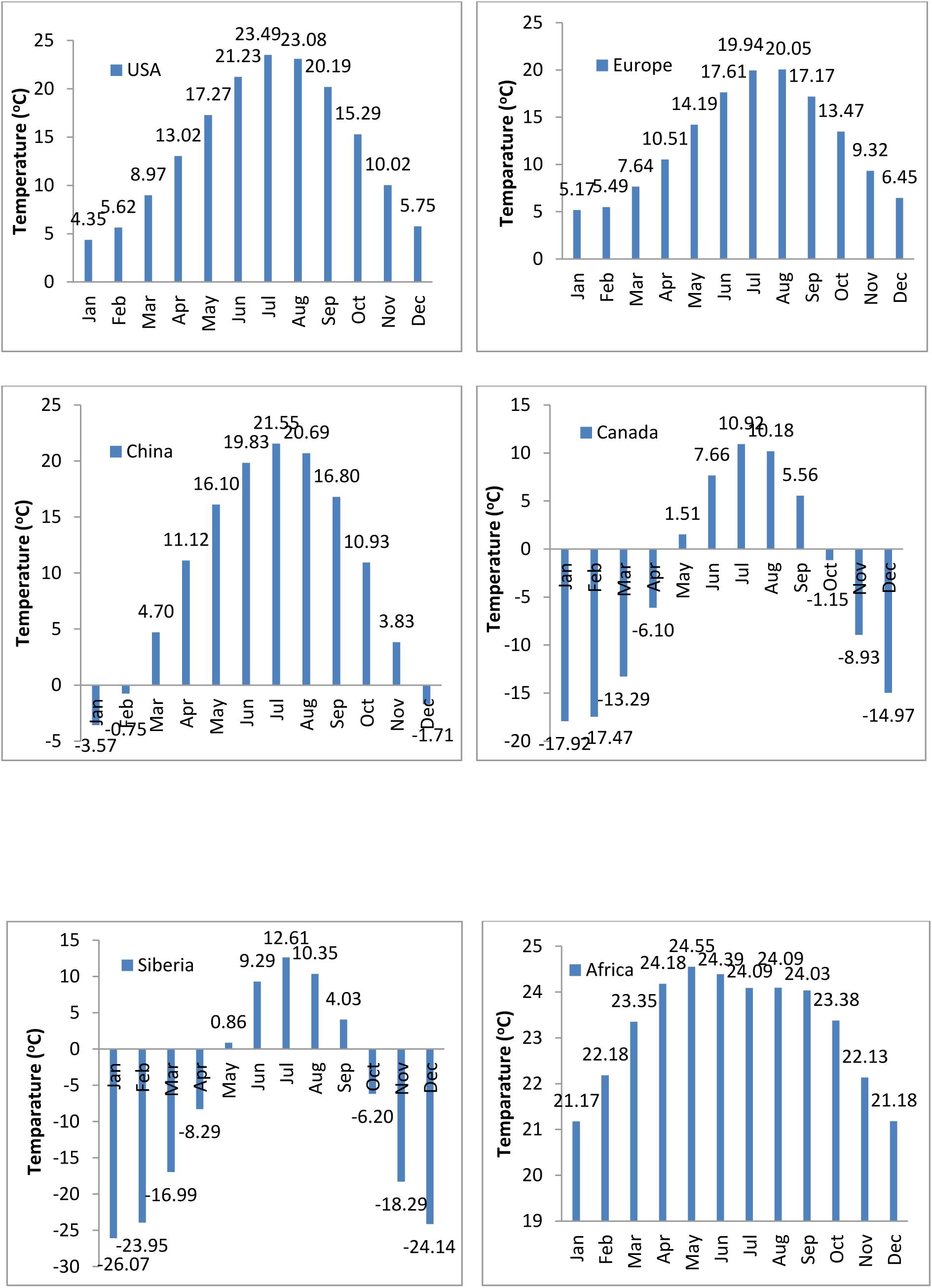
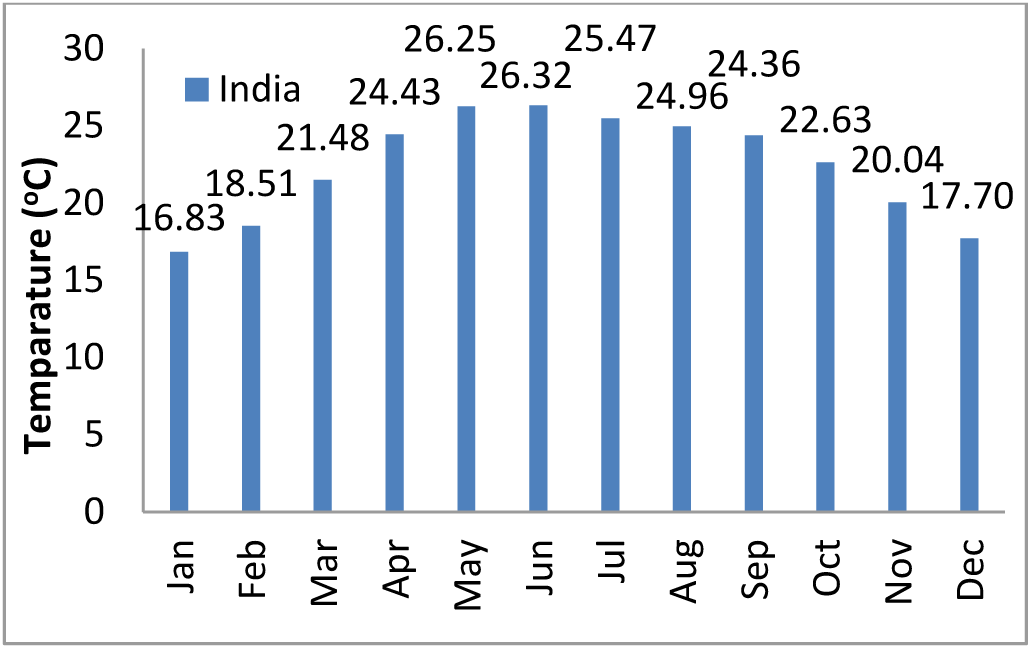
Mean monthly temperatures in the last 41 years (1979–2019) for the highly affected countries a) USA, b) Europe, c) China and least affected countries d) Canada, e) Siberia, f) Africa, g) India.

During the past four months (Nov 2019 - Feb 2020) in the considered regions, temperatures observed are high in November and low in January. However, the highest and lowest temperatures recorded for highly affected regions USA, Europe and China are 10.02°C, 9.32°C, 3.83°C and 4.35°C, 5.17°C, −3.57°C respectively. As well as the least affected areas Canada, Siberia, Africa, India have observed −8.93°C,-18.29°C, 22.18°C, 20.4°C and −17.92°C, −26.07°C, 21.17°C, 16.83°C respectively. Indeed, the highest temperature in Africa is observed in February rather than November. Further, based on the highly affected regions considering past four months temperature data, the favorable temperature (Figure 7) for the growth of Covid-19 is statistically estimated to be between −6.28°C and 14.51°C with a standard deviation of 4.13°C. Therefore, based on this obtained temperature range, it could be assumed that the intensity of Covid-19 in the highly affected regions (USA, China & Europe) would start decreasing from April and substantially disappear by October. Similarly, the pandemic may intensify in Canada between May to October and in Siberia between May to September. Indeed, Africa probably may not get affected largely.

**Figure 7:**
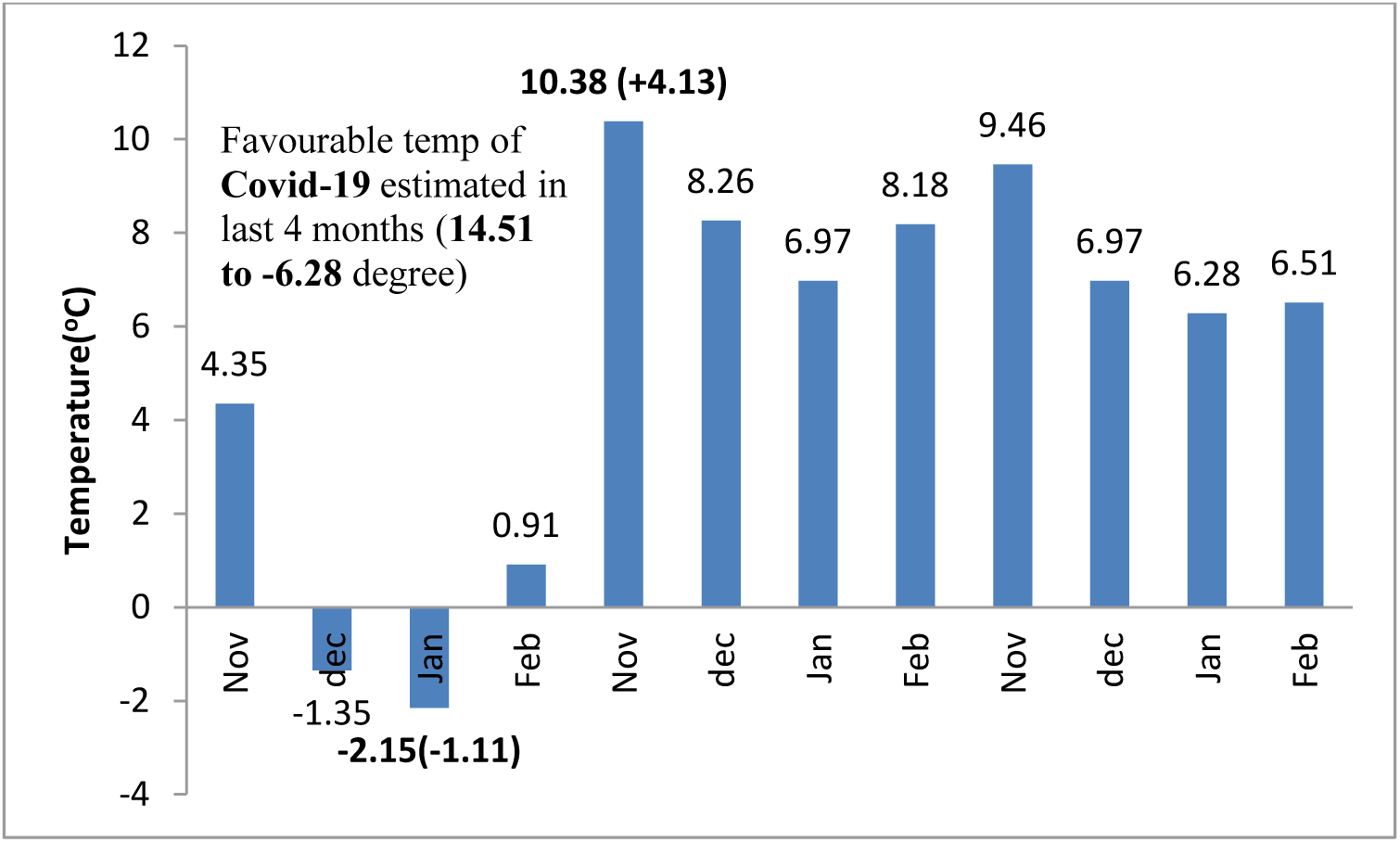
Expected favorable temperature ranges estimated for Covid-19

## Modeling and analysis

An insight into the nature of the global data collected on Covid-19 pandemic shows spatial randomness similar to Poisson’s distribution. Due to discontinuous characteristics or discrete nature of data during the occurrence of infections or events, and could be reasonably applied to estimate the number of events occurring in a particular space. However, it is probably exhibiting independence in time. In this distribution function, events are occurring independently and continuously but without previous memory. Hence, the cumulative distribution/density of this function (CDF) is nothing but a step function. The occurrence of an infection within an area is assumed to follow a Poisson distribution. In estimating hazard rate, the probability of infection (Z) at a given site will exceed a specified level (z), during a specified time (T) such as 14 days, is represented by the following expression:

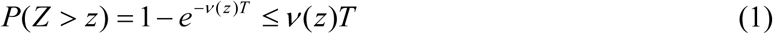

Where *^v^*^(^*^z^*^)^ is the mean per day rate of exceedance of confirmed cases *Z* with respect to *z*. The function *^v^*^(^*^z^*^)^ incorporates the uncertainty in size and location of future events.

## Parameter Estimation

The infection activity of a region could be described based on two parameters such as temperature and cumulative frequency or the rate of occurrences of a particular infection. Gutenberg Richter (1944) developed a relationship which assumes an exponential distribution of magnitude and is expressed as:

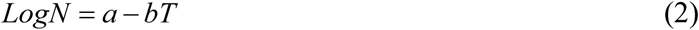

Where ‘a (intercept)’ and ‘b (slope)’ are the constants of regression, that actually describes the infected cases of a region, *N* is the mean daily rate of exceedance of certain particular confirmed cases and T is the temperature. These two parameters could be used as the key inputs in estimating mortality studies. The higher *a*, *b* values indicate a higher level of infection (a-value) with a larger proportion of smaller size events (b-value) or in other words, the b-value describes the relative size distribution of temperature ranges. Indeed, the importance of b-value is such that a small change in b-value results in large changes in occurrences of the projected number of higher temperature dependant infections.

## Results and discussion

The G-R relations for minimum, average and maximum temperature cases affecting the total number of infection cases observed from January 2020 to 4^th^ February 2020 (Wang et al. 2020) are represented in Figure 8. However, from all these figures, when the temperature drops below 0°C, found that the slope of the line is zero, suggesting that the growth rate or infection cases are temperature independent (constant rate). However, as temperature rises greater than 0°C, it shows a significant effect on the decrease in the growth rate of infection. Further, the value of ‘a’ in the G-R relationships of the three cases minimum (2.07), average (3.8456) and maximum (2.5191) temperatures having low, high and median values respectively represent the average temperature considering extreme ranges as the most favorable range for the growth of Covid-19 and therefore, cases would be much higher in this mix condition. However, a lower growth rate is found in the case of minimum temperature ranges and the median growth is observed in case of maximum temperature. Therefore, the average temperature is the highly vulnerable range for Covid-19’s higher growth experienced seasonally by a region or a country. On the other hand, the higher ‘b’ value (0.0725) is found in case of min temperature and the least value (0.0594) is found in case of max temperature whereas median value (0.0613) found in case of avg temperature indicating higher value representing the lesser proportion of higher temperature ranges and vice versa.

**Figure 7:**
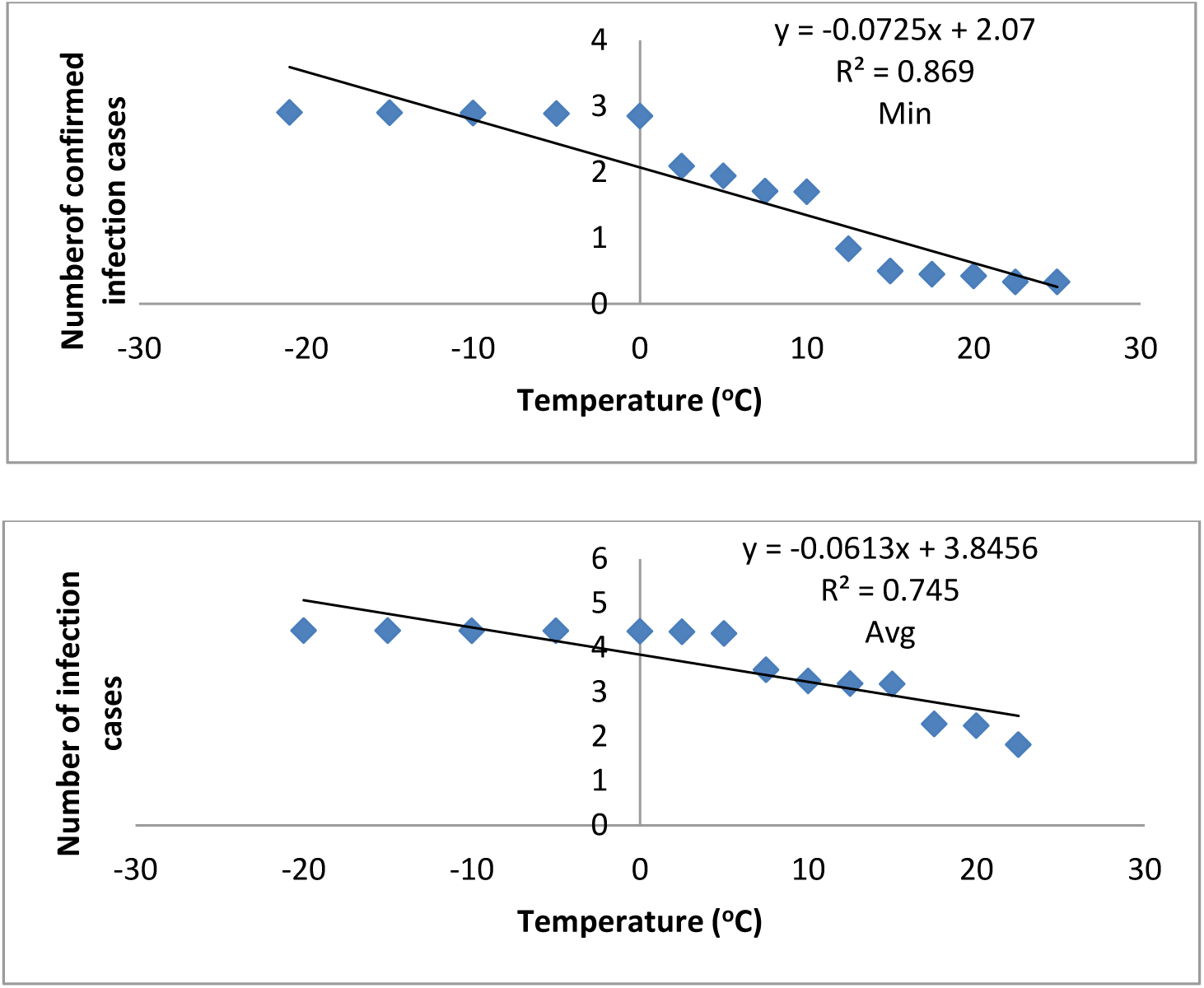
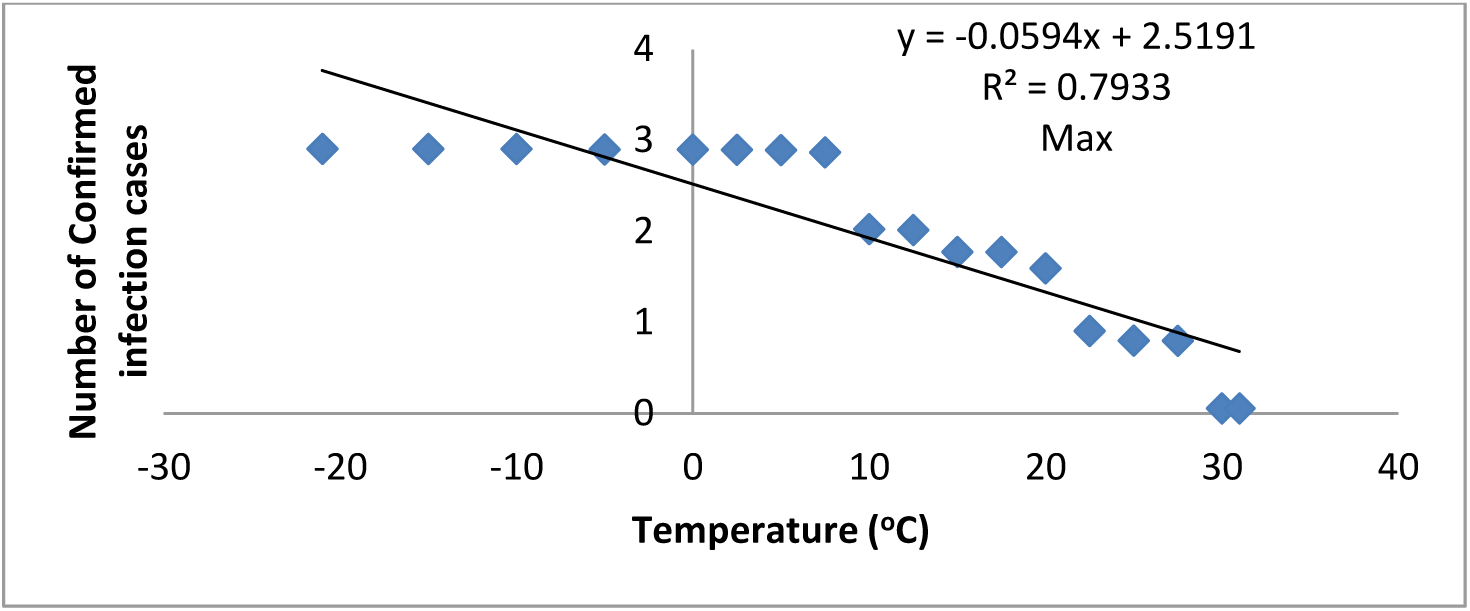
Relationship between the temperature and number of infected cases extracted from the global data during Jan 2020-4^th^ Feb 2020 a) minimum, b) average c) maximum

Accordingly, from these outcomes, the total number of infections due to Covid19 decreases from 13–16 peoples per day, due to 1°C rise or change in temperature, which is a significant sign or impact as a result of the increase in temperatures. As per the present analysis, the strong environmental factor which could retard the growth rate of infection is temperature. Also, it can be expected or believed that the growth the rate may drops down in a reasonable rate for some countries such as India and Australia.

## Conclusions

The growth rate of the global pandemic is related to the changes in temperature and found to be the most influential environmental parameter that could retard the growth by 13–16 new cases/day with a 1°C rise in it. The statistical estimation suggests −6.28°C and 14.51°C, as the most the favorable temperature range for the growth of COVID-19. Also as per the analysis, the intensity of COVID-19 in the highly affected regions (USA, China & Europe) would start decreasing from April and substantially disappear by October. Similarly, the pandemic may intensify in Canada between May to October and in Siberia between May to September. Indeed, Africa probably may not get affected largely.

## Data Availability

all reference and data source given

## Acknowledgement

The outcome of the work in this paper is dedicated to the welfare of the society and nation in particular to the COVID-19 victim.

## Notes

### Competing Interest Statement

authors have no competing interest

### Funding Statement

requested for funding agencies

